# Identifying functional drivers of Hepatoblastoma outcomes via agent-based modeling and transcriptomics

**DOI:** 10.64898/2026.08.07.26359940

**Authors:** Alessandro Ravoni, Yuanhua Liu, Stefano Cairo, Filippo Castiglione, Christine Nardini

## Abstract

Hepatoblastoma (HB) is the most common pediatric liver cancer and represents a major clinical challenge, due to the lack of effective therapies for advanced stages and disease relapse. In this work, we use the results of a previously HB-tailored agent-based model of the immune system to investigate whether model-derived variables can be of use in the prediction of patients’ outcomes. To this aim, we apply factor analysis to the results of a simulated cohort of HB patients, to identify combinations of key immunological variables able to discriminate disease outcomes in the simulator, and we then assess the coherence of such predictions with independent results of differential expression and enrichment analyses on HB transcriptomics. Our analysis proposes that the ability of immune cells, particularly natural killer and CD8^+^ cytotoxic T cells, to recognize tumor-associated antigens and exert cytotoxic activity is essential for disease control following treatment.

## 1 Introduction

Hepatoblastoma (HB) is the most frequent pediatric liver cancer [1, 2]. Molecular and genetic studies have shown that HB can be classified into several subgroups [2]. Among these, the earliest and still well recognized classification [3] defines two main transcriptomic subtypes (C1 and C2), identified using a 16-gene signature. The C1 subtype is mainly associated with fetal histology and positive treatment response (3-year overall survival (OS) and event-free survival (EFS) of 90 *−*95% and 87%, respectively), while the C2 subtype is characterized by a stronger proportion of highly proliferative embryonal tumor cells and associated with significantly reduced survival probabilities (3-year OS and EFS rates of 68% and 63%, respectively) [1].

Recently, we introduced an *in silico* model simulating the combined effects of immune response and standard treatment for HB (surgical resection plus cisplatin-based chemotherapy) [4]. The simulator, based on the agent-based model (ABM) of the immune response C-ImmSim [5], was able to reproduce HB heterogeneity and a wide range of disease outcomes consistent with clinical data, including 3-year OS and EFS estimates. In this current study we explore whether C-ImmSim output at the time of resection can predict patients outcome. Further, we explore whether these features correlate with transcriptomic data. Since transcriptomics was not used to calibrate the model, this provides an initial validation step, supporting model consistency.

In particular, we follow two complementary approaches. First, we apply factor analysis (FA) [6] to identify which simulated immune mechanisms are most strongly associated with the outcomes of HB. Second, we perform differential gene expression analysis on HB data, followed by enrichment analysis, to identify the biological processes characterizing cancer subtypes, finally we explore the coherence of the two approaches. Overall, this provides interesting insights into the interaction between the immune system and HB.

## 2 Data and Methods

### 2.1 Modeling immune response in HB under treatment

Here, we briefly recall the model introduced in [4], which simulates the combined effects of the immune response and treatment in HB. C-ImmSim (https://wwwold.iac.rm.cnr.it/filippo/cimmsim/index.html) is a well-established ABM model designed to simulate the immune response to specific antigens. It reproduces both humoral and cellular immune responses by incorporating key biological processes, including bone marrow cell homeostasis, an extended repertoire, thymus selection, self–nonself discrimination, antigen recognition and processing, clonal selection, antibody hypermutation, and memory formation [5]. The model includes several types of entities, namely white blood cells (WBC) such as T and B lymphocytes, macrophages (MA), dendritic cells (DC), epithelial cells and natural killer cells (NK), and molecular components such as antibodies, antigens, immune complexes, cytokines. The simulated dynamics mimic those of the immune system: agents move within a virtual organ, exchange signals, change phenotype, interact, and execute processes governed by stochastic rules defining their behavior.

In [4], the ABM was extended to reproduce the progression of HB by introducing proliferating cancer cell entities. Each cancer cell carries a tumor-associated antigen and can interact with components of the immune system including CD8^+^ cytotoxic T cells (TC), NK, and, to a lesser extent, antibodies, as observed in clinical settings [7]. In addition, cancer cells are subject to the cytotoxic effects of chemotherapy and can be removed through surgical resection. Upon cell death, antigen processing triggers an immune response, which may or may not be effective in controlling cancer growth. In order to capture the key characteristics of the C1 and C2 HB subtypes, we introduce two types of cancer cells that differ only in their mean duplication times. The HB is classified as C1 if the number of cells that replicate more slowly exceed a threshold at the time of resection, otherwise as C2. The model is able to reproduce key clinical outcome measures, such as 3-years OS and EFS rates, as well as the stratification into C1 and C2 subtypes, within a population of virtual patients. These virtual patients differ in terms of immune complexes, immune repertoire, baseline immune cell counts at homeostasis and the stochastic realization of probabilistic events. For each virtual patient, the model also tracks the populations of immune cells and the concentrations of molecules involved in the immune response.

### 2.2 Factor analysis and classification

FA is a statistical method used to reduce the dimensionality of a dataset consisting of *p* variables (the simulated immunological entities) measured on *n* observations (the virtual patients). It aims to extract a small number *m < p* of latent factors that account for the variability and correlations among the observed variables according to the relation *D* = *SL*^*⊤*^ + *ϵ*. Here *D ∈* ℝ^*n×p*^ is the *data matrix*, where the element *D*_*i,j*_ is the value of the *j*-th variable for the *i*-th observation; *S ∈* ℝ^*n×m*^ is the *factor scores* matrix, representing the coordinates of the *n* observations in the latent factor space; *L ∈* ℝ^*p×m*^ is the *factor loadings* matrix, where *L*_*j,f*_ represents the weight of the *j*-th variable on the *f* -th factor; and *ϵ ∈* ℝ^*n×p*^ represents the residual noise, i.e. the part of the data not explained by the latent factors.

FA is a close relative of PCA, with however a particular focus on the common variance among variables, helpful in molecular data analysis interpretation [8]. We use FA to identify combinations of simulated biological entities that capture the observed correlations, with the aim of recognizing for such combinations a specific immune response process. We then apply standard classification methods to identify regions of the factor space that represent groups of virtual patients in which, ideally, similar immune mechanisms are activated, thereby influencing disease progression. In particular, immunological variables are collected at the clinically relevant time point on the day of resection. Factor analysis was performed using the Python FactorAnalyzer module (v0.5.1), employing a maximum likelihood extraction method with varimax rotation and default parameter settings.

### 2.3 HB data analysis

We downloaded 60 RNAseq samples, including tumor and normal parts of the liver from 24 HB patients, from GEO (GSE104766) [9]. Metadata include RC2 (recurrence) and A (alive) labeling. Preprocessed raw count matrices were used for differential analysis, without additional processing of raw sequencing reads. Differential analysis between RC2 and A was performed using DESeq2 [10], where individual effect nested within groups was considered to identify the RC2 specific features (genes). This was implemented through a design formulated in R as “Groups + Groups:patients + Groups: tissues”, where “Groups” are RC2 and A, “patients” is patient ID, and “tissues” refers to the sources of the samples (tumor or normal). DESeq2 filtered lowly expressed genes, retaining those with at least 5 counts in at least 2 samples. Genes with more than 2 fold-change and less than 0.05 adjusted p-value were identified as significant and further processed to identify the associated relevant biological processes using R package profilercluster and GSEA for functional enrichment of the immune associated genesets (obtained from immport, https://www.immport.org) in preranked mode, where the ranking is obtained from the output of DESeq2. This resulted in 1419 upregulated and 1767 downregulated genes.

## 3 Results

### 3.1 Identification of proxies for outcome prediction

The ABM allows us to follow the dynamics of several immunological variables and events. In particular, we track the number of cancer cells killed by TC and NK cells (output of the simulator), observing that both cell types play a more prominent role in cancer cells killing during the early weeks of treatment in non-progressive patients compared to progressive ones (Figure 1, top panel in the left column). This discrepancy may be mainly driven by two mechanisms: an impaired activation of the immune response, likely due to inefficient antigen processing and presentation following chemotherapy-induced antigen release; and/or reduced cytotoxic efficiency of TC and NK cells against cancer cells, related to suboptimal recognition of tumor associated antigens and altered MHC class I expression.

**Figure 1:**
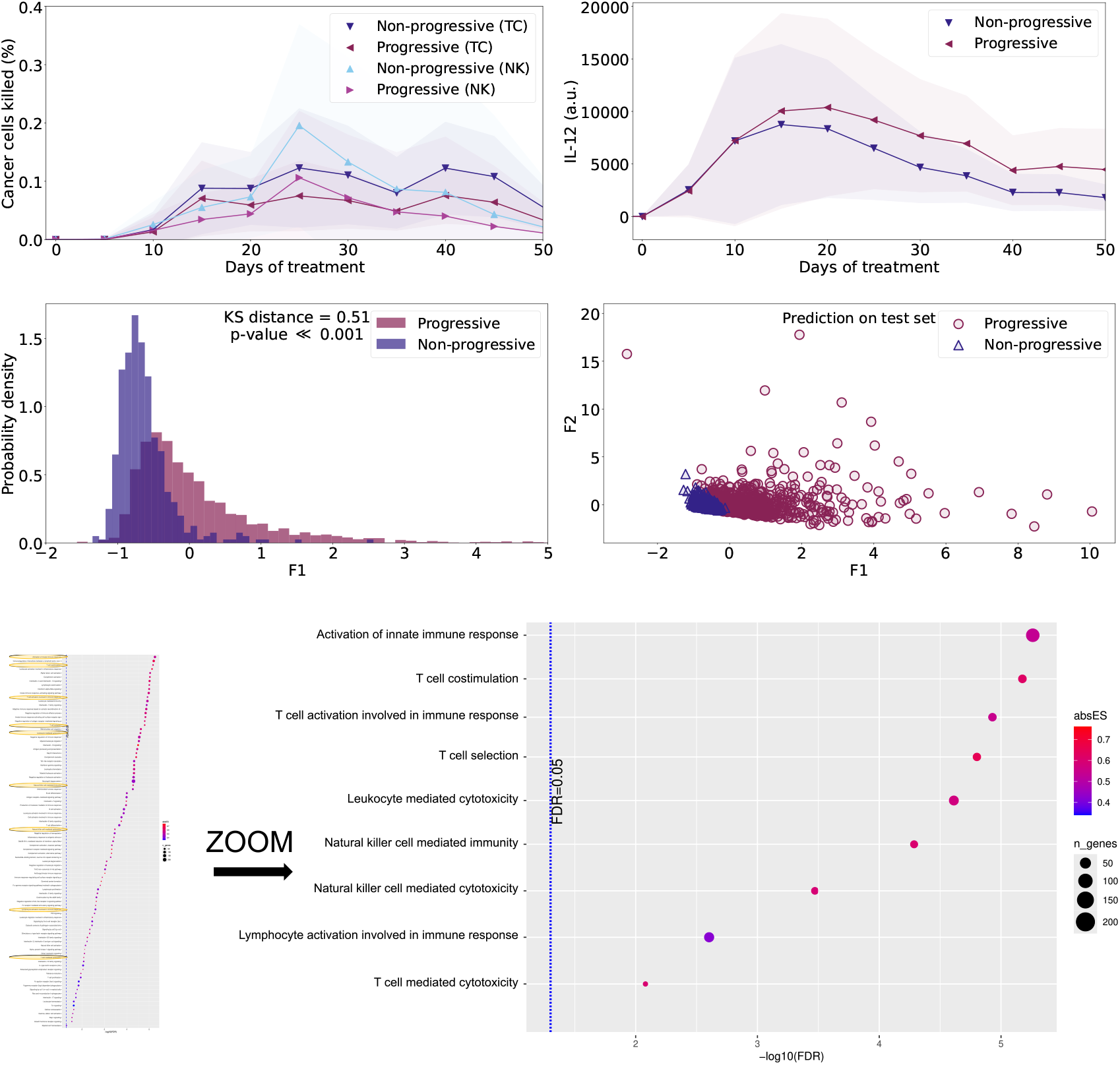
Processes underlying disease outcomes. Top left panel: percentage of cancer cells killed by NK and TC cells in progressive and non-progressive disease during treatment. Top right panel: amount of IL-12 for the two disease outcomes. Middle left panel: distributions of different outcomes with respect to factor F1. Middle right panel: predicted classification of long-term disease in the test set in F1 and F2 space. Bottom panel: significantly downregulated functions in RC2 samples; highlight and zoomed-in: ABM associated functions.

To understand the processes underlying the observed results, we employ FA and identify the subset of variables that most strongly contribute to explaining the model output. Our dataset consists of 5000 virtual patients generated as stochastic realizations of the calibrated ABM, representing different immune system states and histories of stochastic events. The patients are classified as C1 (≈ 3500) and C2 (≈ 1500). We split the dataset in trainig (60%) and test (40%) sets, and apply FA on the training set after checking its reliability (p-value ≪0.001 from Bartlett’s test of sphericity, KMO *>* 0.6 from Kaiser–Meyer–Olkin test). The number of factors must be specified *a priori* and is therefore treated as an input parameter. An initial estimate is obtained using Horn’s parallel analysis, and the final selection is refined by retaining only factors with at least one variable showing absolute loading greater than 0.6, in order to ensure interpretability of the latent structure. The obtained latent factors are shown in Table 1.

**Table 1:** FA latent factors. First column: factors. Second column: entities associated with each factor. Values in parentheses indicate the factor loadings (*>* 0.6). Third column: associated biological processes (ABM simulated).

| Factor | Associated entities(loading) | Associated simulated process |
| --- | --- | --- |
| F1 | IL-12(0.91), IL-18(0.80), presenting DC(0.79), total TC(0.72), presenting MA(0.71), memory TC(0.64) | TH1-mediated immune response |
| F2 | memory B(0.85), total B(0.83), IgM(0.84), IgG1(0.77), memory TH(0.60) | humoral response |
| F3 | total NK(0.92), total TH(0.81), baseline WBC(0.77) | Cellular immune responsiveness |
| F4 | IL-10(0.99), TGF-β(0.99) | Immune Regulation |
| F5 | active TH(0.73) | Adaptive cellular activation |

By construction, we search in the factors the underlying common characteristics associated to the system of interest. In our case, the cytokines IL-12 and IL-18 associated with F1 indicate a cellular immune response, in particular linked to CD4^+^ T helper type 1 (TH1) activation and to antigen processing and presentation via MHC class II molecules by MA, ultimately leading to stimulation of a TC-mediated response. The mechanism of antigen processing and presentation via MHC class II is also involved in the humoral immune response, resulting to the production of IgM and IgG antibodies, as well as in TH activation, captured by F2 and F5. F3 and F4 are related to the immunological state of the virtual patients: factor F3 is associated with an increase in the number of immune cells on the day of resection compared to baseline, while factor F4 captures immune regulation involving anti-inflammatory cytokines IL-10 and TGF-*β*.

These factors appear to be relevant for the classification of the long-term disease outcome, namely progressive (cancer recurrence within 3 years after treatment) and non-progressive disease (no cancer recurrence over the same period). In Figure 1, lower-middle panel in the left column, we present, for the training set, the distribution of the virtual patients outcomes projected on F1, which explains the largest proportion of the variability in the data. Although overlapping, the distributions for progressive and non-progressive patients are statistically significantly distant (Kolmogorov–Smirnov, KS =0.42, *p <* 0.001), and in particular we can observe that positive values of F1 are characteristic of a progressive disease. The bottom left panel in Figure 1 shows the predicted outcomes for the test set in the (F1,F2) space, where F2 is the second factor explaining the variability in the data. A partial separation between classes can be observed in this space. In particular, F1 values above 0 and F2 values above 2, are almost exclusively associated with progressive disease. True labels of the test set give consistent results (data not shown).

It is important to note, however, that these projections offer only a partial representation of the underlying structure, which is more appropriately described in the full 5-dimensional factor space where regions associated with different disease outcomes can be identified. In particular, we trained a classifier with logistic regression for disease outcome, with cross-validated (stratified 5-fold) performance metrics: balanced accuracy of 0.75 *±* 0.01 and an ROC-AUC of 0.83 *±* 0.01. Interestingly, comparable performances are observed on the test set, with balanced accuracy/ROC-AUC of 0.74*/*0.82. We also note that alternative classification approaches, such as support vector machines, random forests, and neural networks, yielded similar results (data not shown). These good predictive performance should be interpreted within the *in silico* framework: although the model was calibrated against clinical observations, these results do not constitute final evidence of comparable predictive performance in real-world patients.

### 3.2 Biological processes underlying disease outcomes

The identified factors collectively capture key biological processes represented in the model that underlie long-term disease outcomes, suggesting that reduced cytotoxic activity of NK and TC cells against cancer cells, rather than impaired activation of the immune response, is responsible for cancer progression.

Progressive disease shows, on average, positive values across all factors, particularly higher values of factor F1 compared to non-progressive disease (Figure 1, lower-middle panel in the left column). The loadings of associated variables are consistently positive (Table 1, right column), indicating a direct relationship between factor values and variables. This suggests successful antigen acquisition and processing in progressive disease, enhanced by chemotherapy-driven antigen release, promoting a pro-inflammatory environment that supports TC and NK cytotoxic activity; this results is confirmed by the higher levels of IL-12 in progressive disease (Figure 1, upper-middle panel in the left column), linked in our model to the promotion of the cytotoxic activity of NK cells and to the antigen presentation by MA and DC, that leads the activation of TC cells. The observed low percentage of cancer cell killing by these cells therefore suggests that an increased ability of cancer cells to specifically evade their cytotoxic activity drives disease progression in our model.

It is interesting to observe these results in light of the functions that appear to be elicited in the transcriptomics of real HB samples. Figure 1, right column, presents the enrichment for statistically significantly downregulated functions in RC2 samples. Approximately 10% of the functions can be associated with the mechanisms described above, namely: *Natural killer cell mediated immunity, Natural killer cell mediated cytotoxicity, Activation of innate immune response, T cell costimulation, T cell activation involved in immune response, T cell selection, Leukocyte mediated cytotoxicity, Lymphocyte activation involved in immune response*, and *T cell mediated cytotoxicity*. Despite the necessary caution in drawing conclusions due to the intrinsic differences between the two analyses, it is noteworthy that the overlap between enriched functions and model-derived processes identified through FA provides initial evidence of biological concordance between the ABM and HB disease progression.

## 4 Conclusion

We take a first step toward the validation of an ABM of the immune response against HB, the most common pediatric liver cancer, with two two complementary approaches: (i) FA on simulated results using a model calibrated on clinical data, and (ii) differential expression and enrichment analyses on HB transcriptomics. Among the results, the coherence observed in the reduced activity of NK cells and the low affinity between TC receptors and antigens presented on MHC class I complexes holds great potential for research in *in silico* trials.

## Data Availability

All data produced in the present study are available upon reasonable request to the authors

## References

[1] Cairo S. et al. A combined clinical and biological risk classification improves prediction of outcome in hepatoblastoma patients. European Journal of Cancer, 141:30–39, ec 2020.

[2] Armengol C., Cairo S., and Kappler R. Bridging molecular basis, prognosis, and treatment of pediatric liver tumors. Hepatoma Research, 2021.

[3] Failli S. et al. Computational drug prediction in hepatoblastoma by integrating pan-cancer transcriptomics with pharmacological response. Hepatology, 80(1):55–68, September 2023.

[4] Ravoni A., Mastrostefano E., Kappler R., Armengol C., Castiglione F., and Nardini C. Mimicking cancer therapy in an agent-based model: The case of hepatoblastoma. Computer Methods and Programs in Biomedicine, 269:108917, sept 2025.

[5] Castiglione F. and Celada F. Immune System Modelling and Simulation. Taylor & Francis Group, 2020.

[6] Joliffe I.T. and Morgan B.J.T. Principal component analysis and exploratory factor analysis. Statistical Methods in Medical Research, 1(1):69–95, March 1992.

[7] Ragone C. et al. Identification and validation of viral antigens sharing sequence and structural homology with tumor-associated antigens (TAAs). Journal for ImmunoTherapy of Cancer, 9(5):e002694, may 2021.

[8] Fronza R., Tramonti M., Atchley W.R., and Nardini C. Joint analysis of transcriptional and post-transcriptional brain tumor data: searching for emergent properties of cellular systems. BMC Bioinformatics, 12(1), March 2011.

[9] Hooks KB. et al. New insights into diagnosis and therapeutic options for proliferative hepatoblastoma. Hepatology, 68(1):89–102, 2018.

[10] Michael I Love, Wolfgang Huber, and Simon Anders. Moderated estimation of fold change and dispersion for rna-seq data with deseq2. Genome Biology, 15(12), December 2014.

